# Cardiovascular Outcomes with GLP-1 Receptor Agonists in Patients with Type 2 Diabetes or Obesity Undergoing Surgical Aortic Valve Replacement

**DOI:** 10.64898/2026.06.02.26354773

**Authors:** Justin Lum, Andrew Jordan, Peter Knight, Kazuhiro Hisamoto

**Affiliations:** Division of Cardiac Surgery, University of Rochester Medical Center, Rochester, NY, USA

**Keywords:** GLP-1 receptor agonist, surgical aortic valve replacement, aortic stenosis, type 2 diabetes mellitus, cardiovascular outcomes

## Abstract

**Background:** Glucagon-like peptide-1 receptor agonists (GLP-1 RAs) have demonstrated cardiovascular benefit in type 2 diabetes and obesity, with recent observational data suggesting favorable associations after transcatheter aortic valve replacement. Whether similar associations exist after surgical aortic valve replacement (SAVR) is unknown.

**Methods:** Retrospective propensity-matched cohort analysis using the TriNetX U.S. Collaborative Network. Adults with type 2 diabetes or obesity (BMI ≥30 kg/m²) undergoing SAVR were categorized by GLP-1 RA exposure (any use within 3 months before through 1 year after SAVR) versus no use. One-to-one matching was performed on 44 covariates. Primary outcomes were 1-year all-cause mortality, heart failure, acute kidney injury, acute myocardial infarction, cerebral infarction, and atrial fibrillation. Sensitivity analyses included 30-day landmark restriction and falsification outcomes.

**Results:** After matching, 1,984 patients were retained per cohort. GLP-1 RA use was associated with lower 1-year risks of all-cause mortality (4.8% vs 10.4%; HR, 0.44; 95% CI, 0.34–0.56), acute kidney injury (6.9% vs 10.1%; HR, 0.65; 95% CI, 0.49–0.85), myocardial infarction (3.0% vs 5.1%; HR, 0.57; 95% CI, (0.40–0.82), heart failure (11.3% vs 15.7%; HR, 0.68; 95% CI, (0.51–0.90), and atrial fibrillation or flutter (10.1% vs 13.9%; HR, 0.69; 95% CI, 0.54–0.90; all P≤.006). Cerebral infarction did not differ. In landmark analysis, mortality, heart failure, and acute kidney injury associations persisted; myocardial infarction and atrial fibrillation associations were attenuated. Falsification outcomes were null.

**Conclusions:** Perioperative GLP-1 RA use was associated with lower 1-year cardiovascular event rates after SAVR. These hypothesis-generating findings support prospective randomized investigation.

**Clinical Perspective:** *What Is New?:*

– This is the first analysis to evaluate the association between GLP-1 receptor agonist use and cardiovascular outcomes specifically in patients undergoing surgical aortic valve replacement, extending prior observations from the transcatheter aortic valve replacement literature to a distinct surgical population.
– Perioperative GLP-1 receptor agonist use was associated with significantly lower 1-year risks of all-cause mortality, heart failure, and acute kidney injury, and these associations persisted after 30-day landmark restriction. Associations with acute myocardial infarction and new-onset atrial fibrillation were attenuated to null after landmark restriction, indicating these signals were predominantly driven by peri-procedural events.

*What Are the Clinical Implications?:*

– These hypothesis-generating findings suggest that GLP-1 receptor agonists may have a role as adjunctive perioperative cardioprotection in patients with type 2 diabetes or obesity undergoing surgical aortic valve replacement, though the observed effect magnitudes exceed those from randomized trials and residual confounding cannot be excluded.
– Prospective randomized investigation is needed to determine whether GLP-1 receptor agonist therapy confers genuine cardiovascular benefit in this population and to identify optimal timing of initiation relative to surgery.

## Introduction

Calcific aortic stenosis is the most common valvular heart disease in the United States, affecting 1% to 2% of adults older than 65 years and approximately 12% of those older than 75 years.^1^ Although transcatheter aortic valve replacement (TAVR) is now preferred for many elderly and high-risk patients, surgical aortic valve replacement (SAVR) remains the standard of care for younger patients, those with bicuspid anatomy, those requiring concomitant cardiac procedures, and those with anatomy unsuitable for transcatheter approaches.^2^ Despite improvements in operative technique, post-SAVR cardiovascular morbidity and mortality remain substantial: contemporary national data demonstrate 1-year mortality of 2.6% even in low-risk patients, with higher rates in those with impaired ventricular function, renal dysfunction, or diabetes.^3,4^

Type 2 diabetes mellitus and obesity, which are both highly prevalent in the SAVR population, are independently associated with adverse postoperative outcomes. Diabetes confers excess long-term mortality after isolated SAVR (HR, 1.39; 95% CI, 1.03-1.86), with insulin-treated patients at particularly elevated risk.^5^ Obesity increases the incidence of acute kidney injury, sternal wound infection, and resource utilization after SAVR.^6^ Acute kidney injury complicates 12% to 31% of SAVR procedures and is associated with prolonged intensive care stays and increased short- and long-term mortality.^7^ New-onset atrial fibrillation occurs in 37% to 51% of patients after surgical aortic valve replacement and independently predicts subsequent arrhythmia recurrence and death.^8,9^

Glucagon-like peptide-1 receptor agonists (GLP-1 RAs) have demonstrated cardiovascular benefit in randomized trials of patients with type 2 diabetes (LEADER, SUSTAIN-6, REWIND)^10–12^ and obesity without diabetes (SELECT).^13^ Proposed mechanisms include attenuation of systemic inflammation, improved endothelial function and nitric oxide bioavailability, plaque stabilization, weight reduction, blood pressure lowering, and natriuretic effects.^14,15^

Recent observational analyses have reported favorable associations between GLP-1 RA use and cardiovascular outcomes after TAVR.^16,17^ Whether similar associations exist after SAVR is unknown. The SAVR population differs from TAVR cohorts in age, baseline surgical risk, life expectancy, anatomic complexity, and recovery physiology, with the latter involving cardiopulmonary bypass and median sternotomy, making extrapolation from TAVR data uncertain. We conducted a retrospective propensity-matched cohort analysis to evaluate the association between perioperative GLP-1 RA use and 1-year cardiovascular outcomes in adults with type 2 diabetes or obesity undergoing SAVR.

## Methods

### Study Oversight

This study was conducted in accordance with the Strengthening the Reporting of Observational Studies in Epidemiology (STROBE) guidelines for cohort studies. Because the analysis used aggregated, de-identified data from a federated research network, it was exempt from institutional review board approval under 45 CFR §46.104(d)(4).

### Data Source

We used the TriNetX Analytics Network (TriNetX, Cambridge, MA), a multicenter federated research platform that aggregates de-identified electronic health record (EHR) data from more than 275 million patients across over 120 healthcare organizations.^18^ Analyses were conducted within the U.S. Collaborative Network, which comprises more than 100 million patients across 73 responding healthcare organizations. The platform provides patient-level analytics including cohort creation, propensity score matching, time-to-event analysis, and risk comparisons across matched cohorts. All queries were executed on May 20, 2026.

### Study Population and Design

We performed a retrospective propensity-matched cohort analysis of adults aged 18 years or older with a documented diagnosis of type 2 diabetes mellitus (ICD-10-CM E11) or overweight and obesity (ICD-10-CM E66) who underwent SAVR. Procedures were identified using CPT codes 33405, 33406, 33410, 33411, and 33412 and ICD-10-PCS codes 02RF07Z, 02RF08Z, 02RF0JZ, and 02RF0KZ. Both isolated SAVR and SAVR with concomitant procedures were included. Ross procedures (CPT 33413), which constitute a small minority of adult SAVR procedures and involve concurrent pulmonary valve replacement with distinct physiology, were excluded from both cohorts. The SAVR procedure served as the index event for both cohorts.

Patients were categorized into two cohorts based on GLP-1 RA exposure. Cohort 1 (GLP-1 RA) included patients with documentation of any of the five GLP-1 RAs with established cardiovascular outcomes data (tirzepatide, semaglutide, liraglutide, dulaglutide, or lixisenatide) within 3 months before, through 1 year after the index SAVR. Cohort 2 (non-GLP-1 RA) included patients with no documented GLP-1 RA use at any time. The 3-month pre-to 1-year post-procedure exposure window was selected to capture realistic perioperative and early outpatient prescribing patterns, including both prevalent users maintained on therapy through surgery and patients initiated postoperatively, consistent with the approach used in prior TAVR analyses.^16^

### Outcomes

Prespecified primary outcomes were all-cause mortality, acute kidney injury (ICD-10-CM N17), acute myocardial infarction (I21), cerebral infarction (I63), heart failure (I50), and new-onset atrial fibrillation or flutter (I48). Outcomes were ascertained over a follow-up period, beginning 1 day after the index event and ending 365 days after the index event. For each outcome, patients with the respective diagnosis documented prior to the follow-up window were excluded from that analysis to ensure an at-risk denominator.

Two prespecified negative-control (falsification) outcomes were included: dorsalgia (ICD-10-CM M54) and osteoarthritis (ICD-10-CM M15-M19). Dorsalgia has no plausible causal relationship with GLP-1 RA exposure. Osteoarthritis has a potential indirect causal pathway through GLP-1 RA-mediated weight loss reducing joint loading, and the SELECT trial reported numerical reductions in osteoarthritis-related symptoms with semaglutide.^13^ Null associations with both negative-control outcomes would argue against gross unmeasured confounding from healthy-user bias.

### Propensity Score Matching

One-to-one propensity score matching was performed using the TriNetX built-in greedy nearest-neighbor algorithm with a caliper of 0.1 pooled standard deviations. Matching covariates included demographics (age at index, sex, race, ethnicity), comorbidities (type 2 diabetes mellitus, overweight and obesity, essential hypertension, ischemic heart disease, heart failure, atrial fibrillation and flutter, chronic lower respiratory diseases, chronic kidney disease), frailty proxies (age-related physical debility, abnormalities of gait and mobility, cognitive symptoms, cachexia), behavioral factors (tobacco use, alcohol-related disorders), concomitant medications (antilipemic agents, antiarrhythmics, beta-blockers, diuretics, ACE inhibitors, angiotensin II receptor blockers, potassium-sparing diuretics, anticoagulants, insulin, oral hypoglycemic agents, SGLT2 inhibitors), and laboratory values (body mass index, hemoglobin A1c, serum creatinine, estimated glomerular filtration rate). A standardized mean difference (SMD) of less than 0.10 was considered indicative of adequate balance.

### Statistical Analysis

Continuous variables are reported as mean ± standard deviation and were compared using independent-sample t-tests. Categorical variables are reported as counts and percentages and were compared using chi-square tests. Time-to-event analyses used Kaplan-Meier estimation with log-rank testing and Cox proportional hazards regression to estimate hazard ratios (HRs) with 95% confidence intervals (CIs). Patients were censored at the last documented encounter within the 365-day follow-up window. The proportional hazards assumption was assessed visually using Kaplan-Meier curves and statistically using the test of proportionality available within the TriNetX platform. Statistical significance was defined as a two-sided P value less than .05.

E-values were calculated for each outcome using the method of VanderWeele and Ding^19^ to quantify the minimum strength of association on the risk-ratio scale that an unmeasured confounder would need to have with both GLP-1 RA exposure and the outcome, conditional on measured covariates, to fully explain away the observed association.

Deaths during follow-up were censored rather than treated as competing events for non-mortality outcomes, as Fine-Gray subdistribution hazard modeling is not natively supported within the TriNetX analytical platform. The direction and potential magnitude of this limitation are addressed in the Discussion.

### Data Access and Responsibility

Justin Lum had full access to all the data in the study and takes responsibility for the integrity of the data and the accuracy of the data analysis.

### Sensitivity Analyses

Two prespecified sensitivity analyses were conducted.

First, a 30-day landmark analysis excluded all outcomes occurring within the first 30 days after the index event, with follow-up assessed from day 31 through day 365. This analysis was designed to mitigate residual immortal-time bias and to separate peri-procedural events directly attributable to the surgical procedure from post-recovery drug effects.

Second, E-values were calculated for both primary and landmark analyses to assess the robustness of each association to potential unmeasured confounding.

## Results

### Patient Selection and Baseline Characteristics

A total of 2,070 patients met criteria for Cohort 1 (GLP-1 RA use within the perioperative window) and 41,532 patients met criteria for Cohort 2 (no GLP-1 RA exposure). After 1:1 propensity score matching, 1,984 patients were retained in each cohort (3,968 total) and were included in the final outcome analyses.

Prior to matching, GLP-1 RA users were younger than non-users (mean age, 66.4 ± 10.3 vs 72.5 ± 12.9 years; SMD, 0.52) and had a substantially higher burden of cardiometabolic comorbidities, including type 2 diabetes mellitus (83.1% vs 46.1%; SMD, 0.84), overweight and obesity (69.6% vs 38.5%; SMD, 0.66), ischemic heart disease, hypertension, heart failure, and chronic kidney disease. GLP-1 RA users also had higher baseline body mass index (34.5 ± 7.1 vs 31.7 ± 6.5 kg/m²; SMD, 0.42) and hemoglobin A1c (7.1% ± 1.6% vs 6.2% ± 1.3%; SMD, 0.62). Use of cardiovascular and antidiabetic medications was uniformly higher in the GLP-1 RA cohort, including SGLT2 inhibitors (36.5% vs 4.3%; SMD, 0.87).

After propensity-score matching, the two cohorts were well balanced across demographic characteristics, comorbidities, frailty proxies, and medication classes, with SMDs below 0.10 for most covariates (Table 1). Matched prevalences were similar for type 2 diabetes (82.5% vs 83.3%), overweight and obesity (68.4% vs 67.9%), heart failure (56.0% vs 55.6%), atrial fibrillation (41.8% vs 40.1%), chronic kidney disease (29.9% vs 29.9%), and SGLT2 inhibitor use (34.6% vs 32.9%; SMD, 0.04).

**Table 1.**
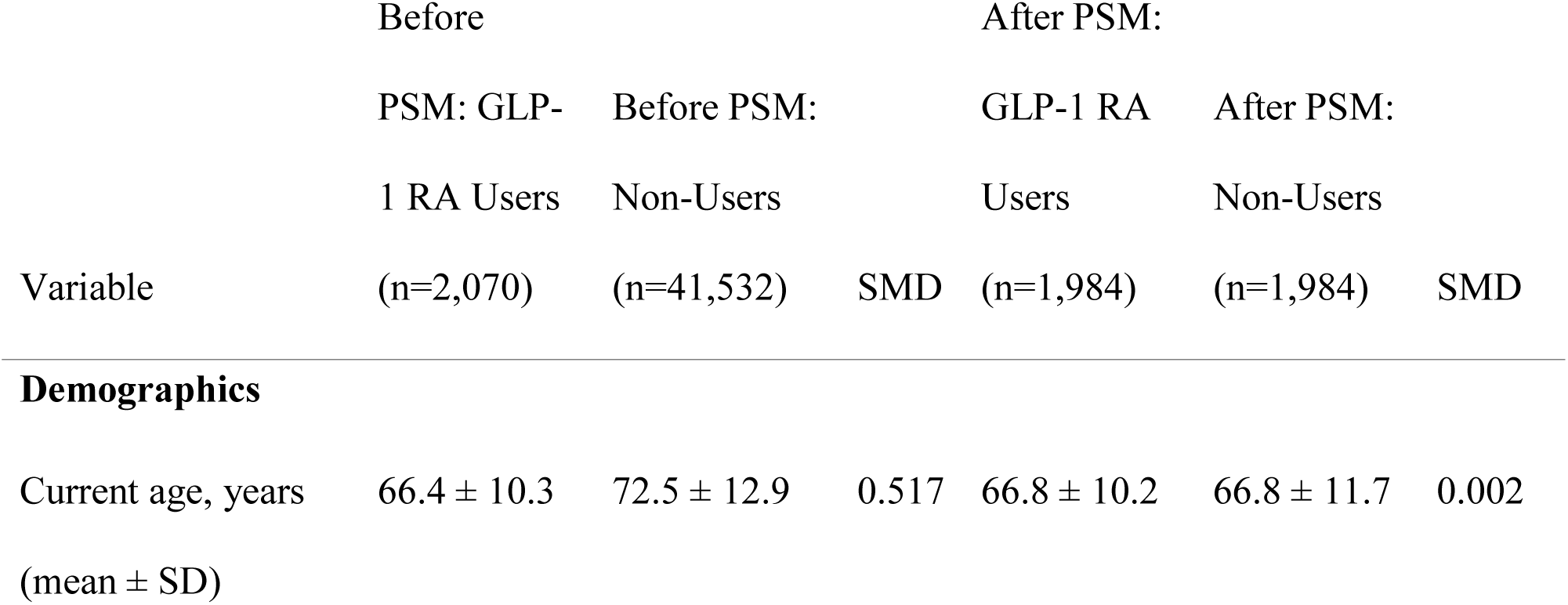
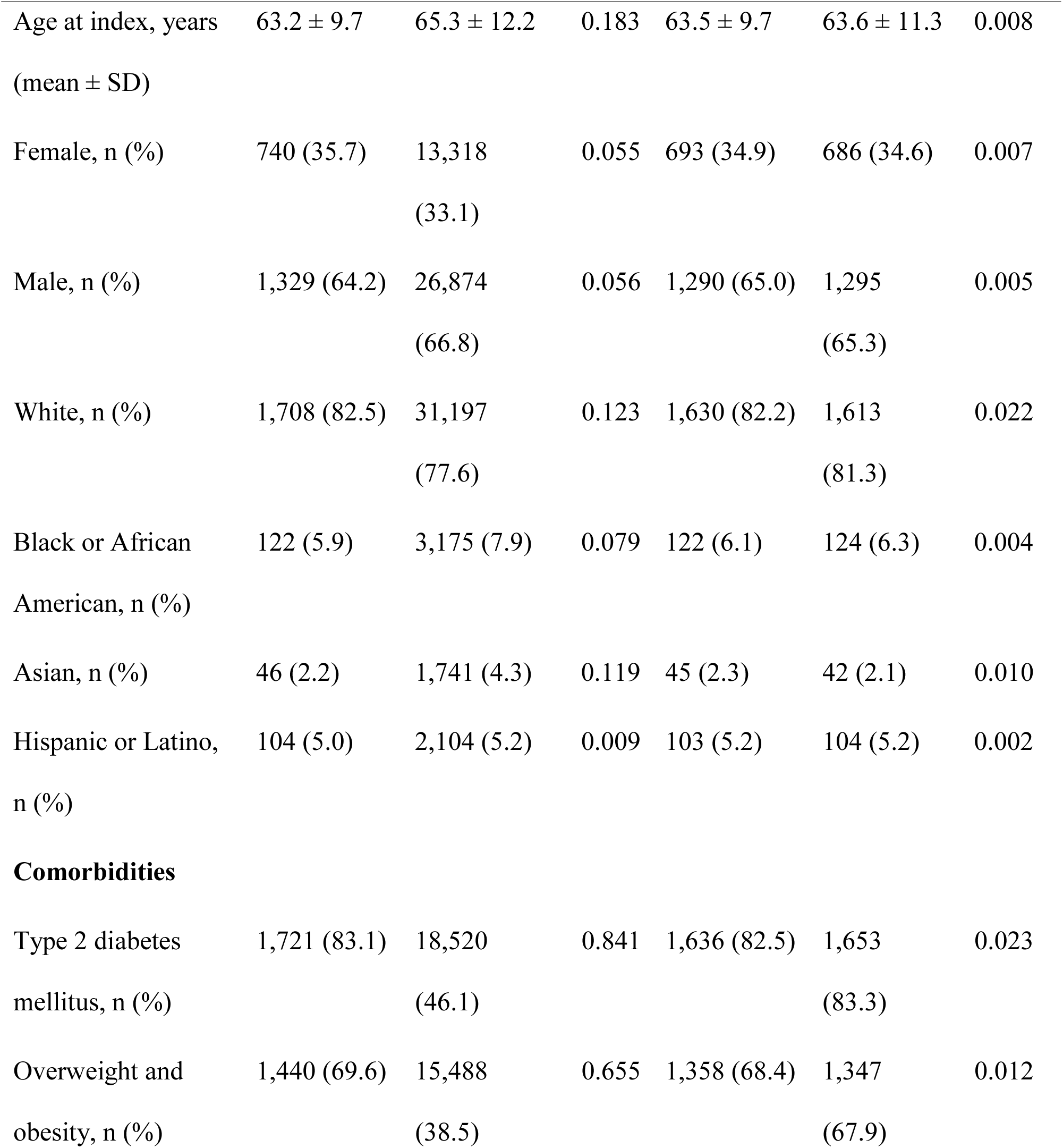
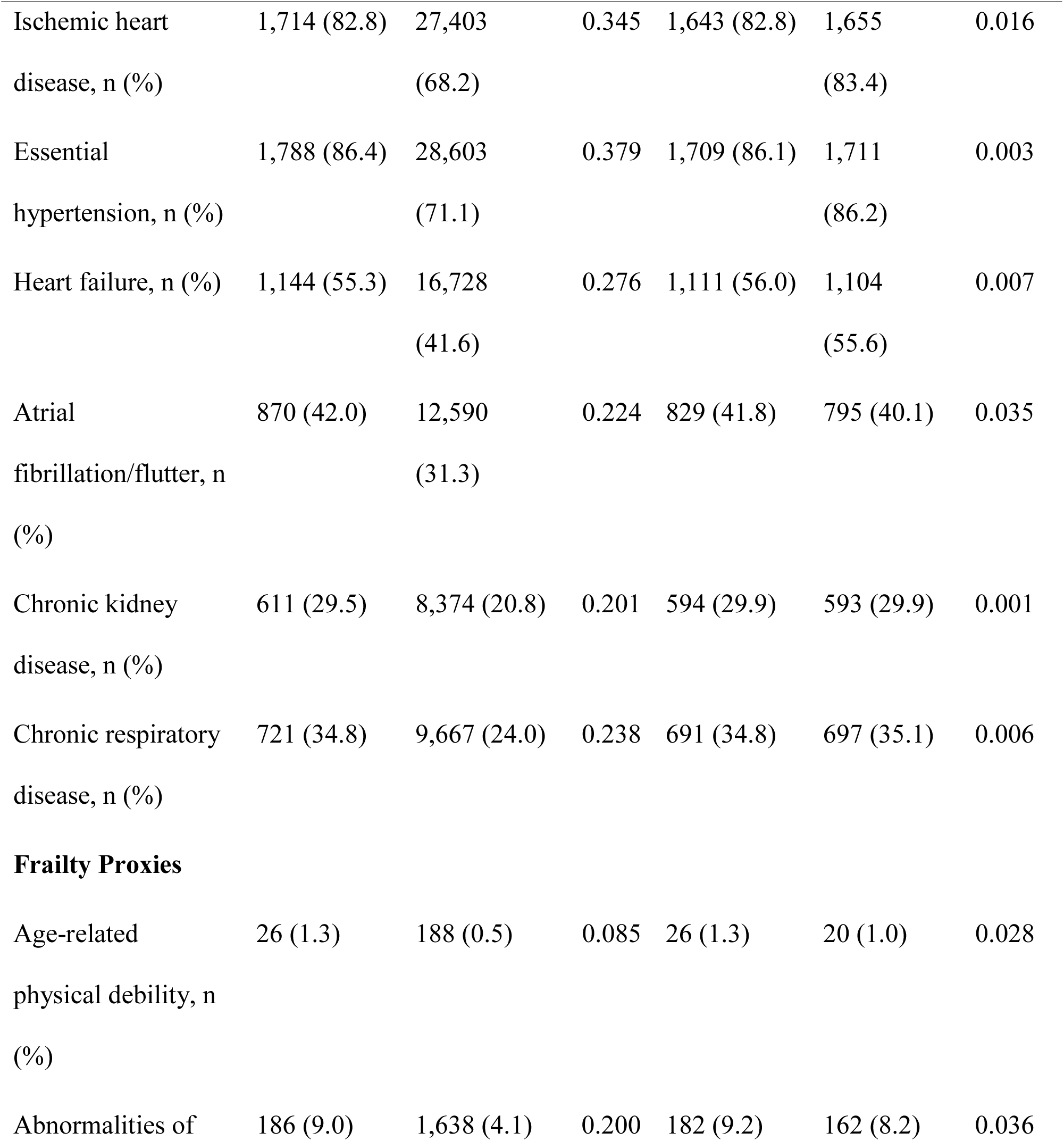
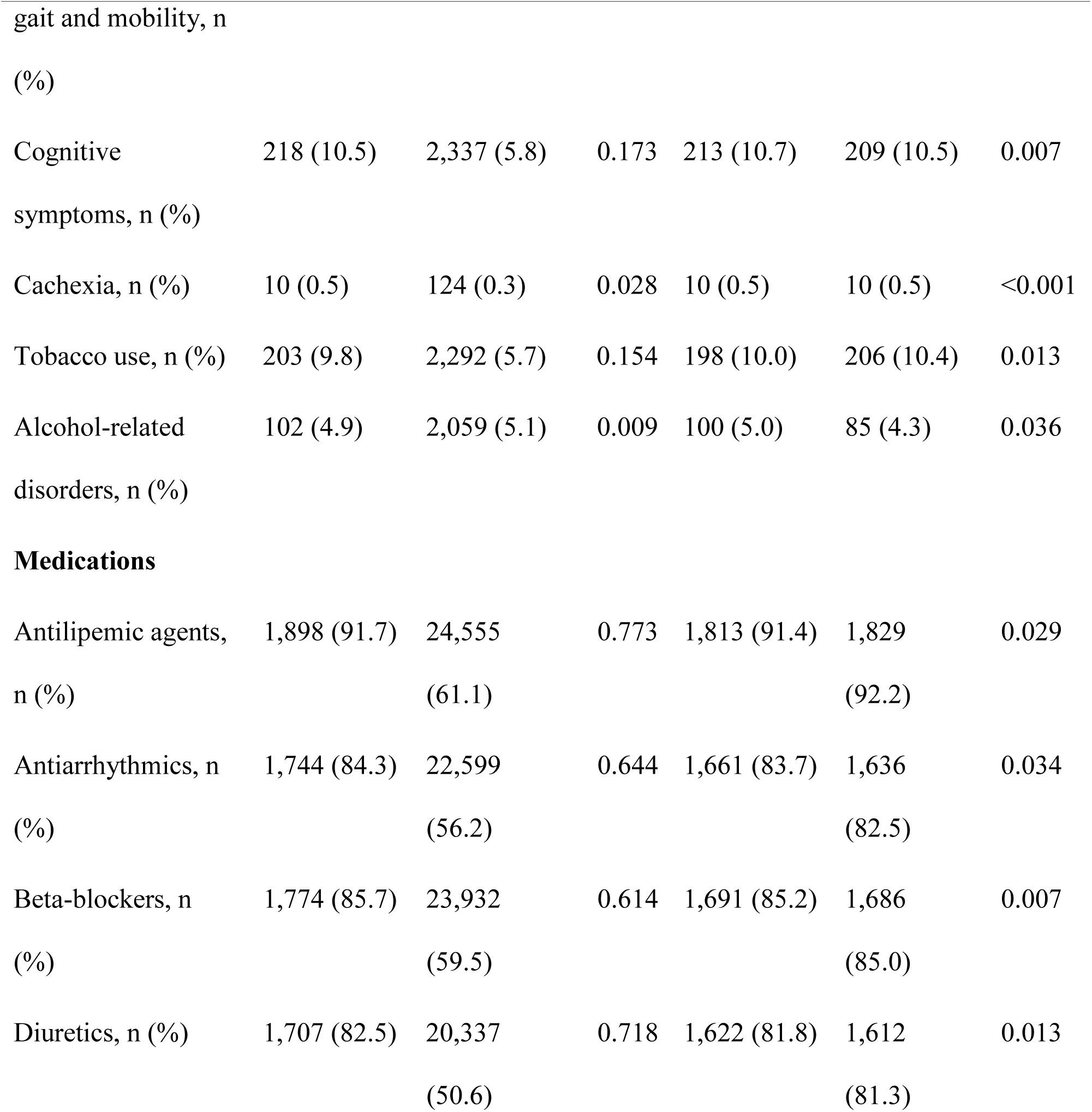
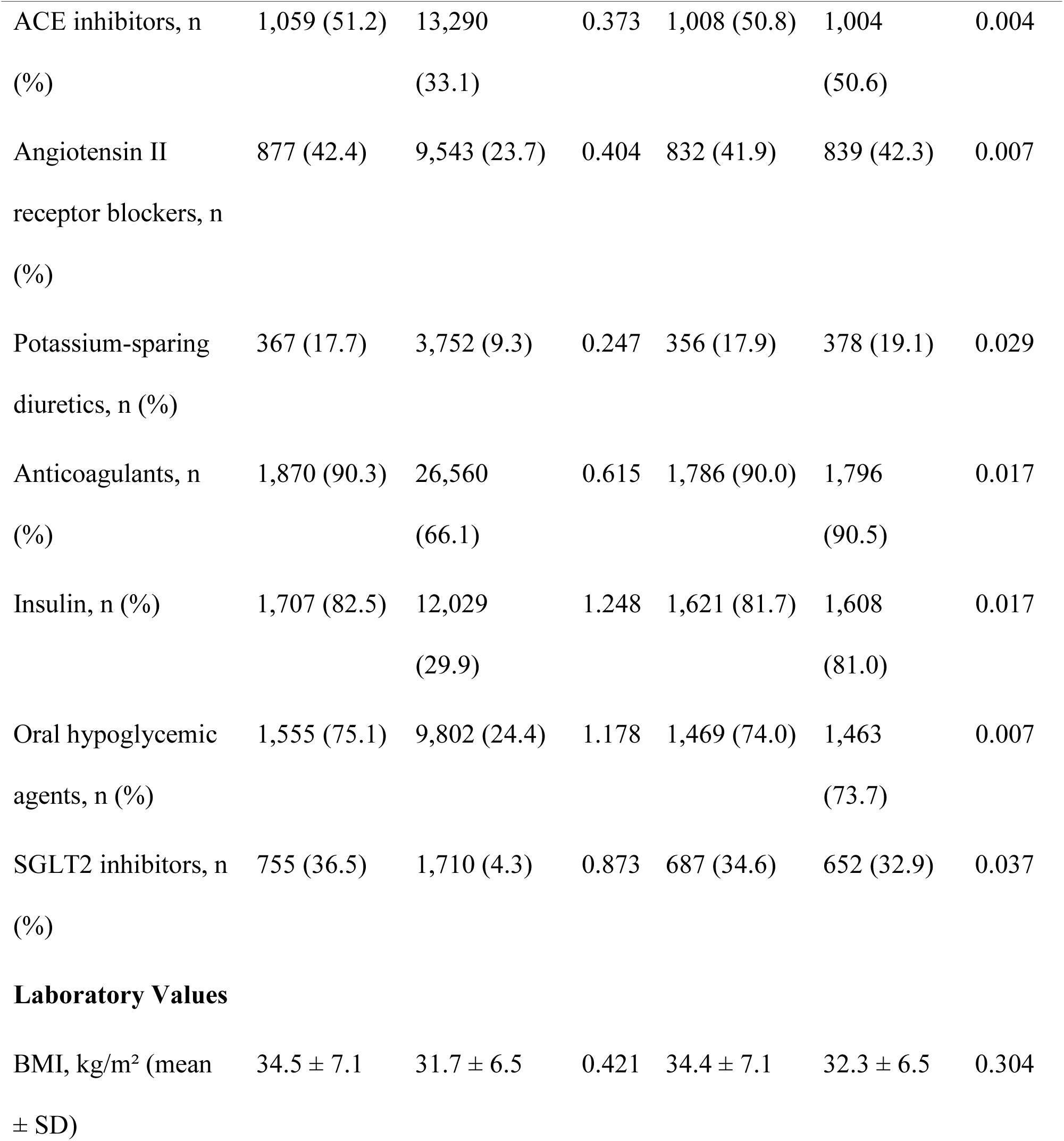
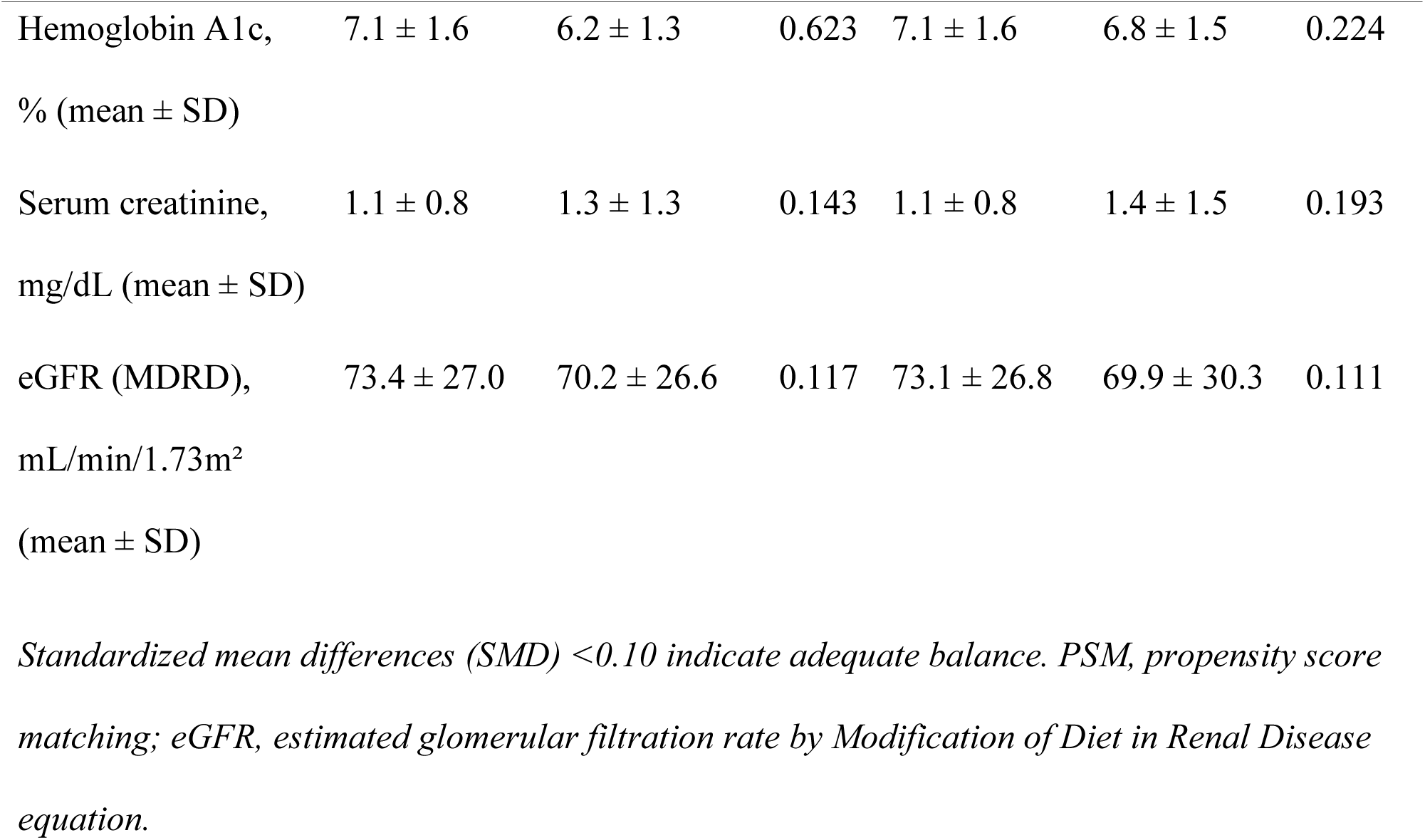
Baseline Characteristics Before and After Propensity Score Matching.

Four covariates exceeded the prespecified SMD threshold of 0.10 after matching: body mass index (34.4 ± 7.1 vs 32.3 ± 6.5 kg/m²; SMD, 0.30), hemoglobin A1c (7.1% ± 1.6% vs 6.8% ± 1.5%; SMD, 0.22), serum creatinine (1.1 ± 0.8 vs 1.4 ± 1.5 mg/dL; SMD, 0.19), and estimated glomerular filtration rate (73.1 ± 26.8 vs 69.9 ± 30.3 mL/min/1.73 m²; SMD, 0.11). The BMI and HbA1c imbalances reflect confounding by indication: these patients were prescribed GLP-1 RAs precisely because of these values, and the residual differences likely capture broader differences in cardiometabolic care intensity that propensity matching on individual covariates cannot fully address. The creatinine and eGFR imbalances favor the GLP-1 RA group with better kidney function at baseline.

### Primary Outcomes

At 1 year of follow-up, GLP-1 RA use was associated with significantly lower risks of all-cause mortality, acute kidney injury, acute myocardial infarction, heart failure, and atrial fibrillation or flutter (Table 2; Figure 1). All-cause mortality occurred in 95 of 1,971 GLP-1 RA users (4.8%) compared with 204 of 1,963 non-users (10.4%) (HR, 0.44; 95% CI, 0.35-0.56; P<.001). Acute kidney injury occurred in 89 of 1,297 (6.9%) versus 118 of 1,166 (10.1%) (HR, 0.65; 95% CI, 0.49-0.85; P<.001). Acute myocardial infarction occurred in 48 of 1,579 (3.0%) versus 72 of 1,408 (5.1%) (HR, 0.57; 95% CI, 0.40-0.82; P=.010). Heart failure occurred in 85 of 750 (11.3%) versus 111 of 707 (15.7%) (HR, 0.68; 95% CI, 0.51-0.90; P=.008). Atrial fibrillation or flutter occurred in 102 of 1,004 (10.2%) versus 135 of 969 (13.9%) (HR, 0.69; 95% CI, 0.54-0.90; P=.005). Cerebral infarction did not differ significantly between cohorts (3.5% vs 4.0%; HR, 0.82; 95% CI, 0.58-1.15; P=.25). The proportional hazards assumption was satisfied for all primary outcomes except acute myocardial infarction, for which the test of proportionality was significant (P=.010); the constant-HR estimate for this outcome should therefore be interpreted with caution.

**Figure 1.**
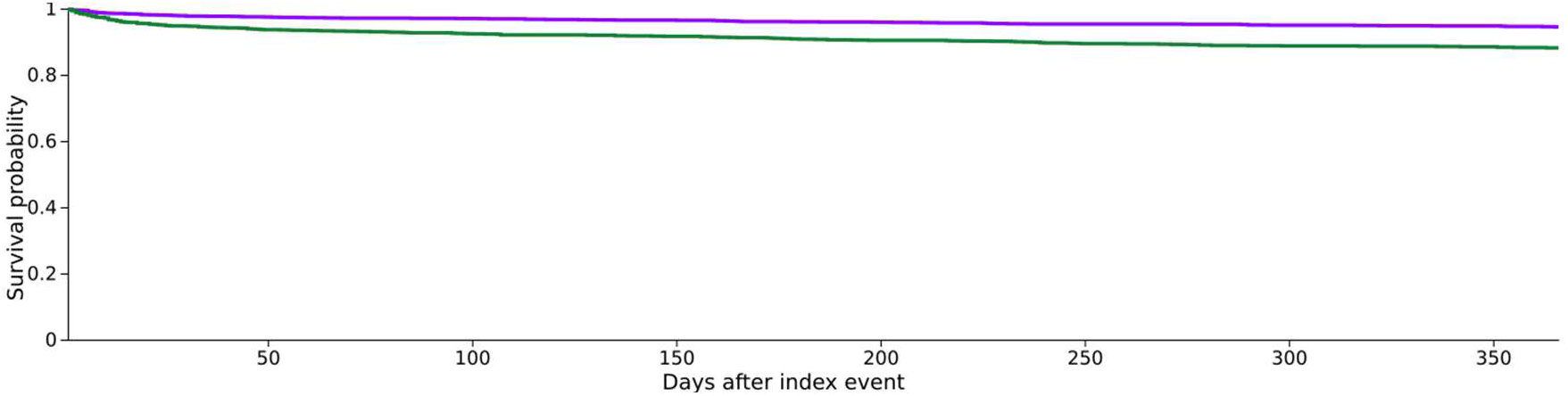
Kaplan-Meier curve for all-cause mortality over 1-year follow-up after propensity score matching. Purple curve represents GLP-1 RA users; green curve represents non-users.

**Table 2.** Primary 1-Year Outcomes After Propensity Score Matching.

| Outcome | GLP-1 RA |  | Hazard Ratio (95% CI) | P Value |
| --- | --- | --- | --- | --- |
|  | Usersevents / at-risk (%) | Non-Usersevents / at-risk (%) |  |  |
| All-cause mortality | 95 / 1,971 (4.8) | 204 / 1,963 (10.4) | 0.44 (0.35-0.56) | <.001 |

| Outcome | GLP-1 RA |  | Hazard Ratio<br>(95% CI) | P<br>Value |
| --- | --- | --- | --- | --- |
|  | Usersevents / at-risk<br>(%) | Non-Usersevents /<br>at-risk (%) |  |  |
| Acute kidney injury | 89 / 1,297 (6.9) | 118 / 1,166 (10.1) | 0.65 (0.49-0.85) | <.001 |
| Acute myocardial<br>infarction | 48 / 1,579 (3.0) | 72 / 1,408 (5.1) | 0.57 (0.40-0.82) | .010 |
| Heart failure | 85 / 750 (11.3) | 111 / 707 (15.7) | 0.68 (0.51-0.90) | .008 |
| Atrial fibrillation/flutter | 102 / 1,004 (10.2) | 135 / 969 (13.9) | 0.69 (0.54-0.90) | .005 |
| Cerebral infarction | 61 / 1,757 (3.5) | 69 / 1,704 (4.0) | 0.82 (0.58-1.15) | .25 |
| Dorsalgia (negative<br>control) | 66 / 1,217 (5.4) | 75 / 1,267 (5.9) | 0.83 (0.60-1.16) | .27 |
| Osteoarthritis (negative<br>control) | 24 / 1,762 (1.4) | 23 / 1,797 (1.3) | 0.99 (0.56-1.75) | .97 |

*Denominators reflect patients remaining after exclusion of those with the outcome documented prior to the 1-day post-index time window. Follow-up was capped at 365 days.*

Both prespecified negative-control outcomes demonstrated no significant association with GLP-1 RA use. Dorsalgia occurred in 66 of 1,217 (5.4%) versus 75 of 1,267 (5.9%) (HR, 0.83; 95% CI, 0.60-1.16; P=.27). Osteoarthritis occurred in 24 of 1,762 (1.4%) versus 23 of 1,797 (1.3%) (HR, 0.99; 95% CI, 0.56-1.75; P=.97). The null associations across both negative-control outcomes argue against gross unmeasured confounding from healthy-user bias.

### E-Value Sensitivity Analysis

E-values for the primary outcomes are presented in Table 3. The E-value for the mortality association was 3.95 at the point estimate and 2.94 at the confidence interval bound, indicating that an unmeasured confounder would need to be associated with both GLP-1 RA exposure and mortality by a risk ratio of at least 2.94 to fully explain the observed association. E-values at the confidence interval bound exceeded 1.4 for mortality, acute kidney injury, acute myocardial infarction, heart failure, and atrial fibrillation. E-values for cerebral infarction and both negative-control outcomes were 1.00 at the confidence interval bound, consistent with their non-significant primary findings.

**Table 3.**
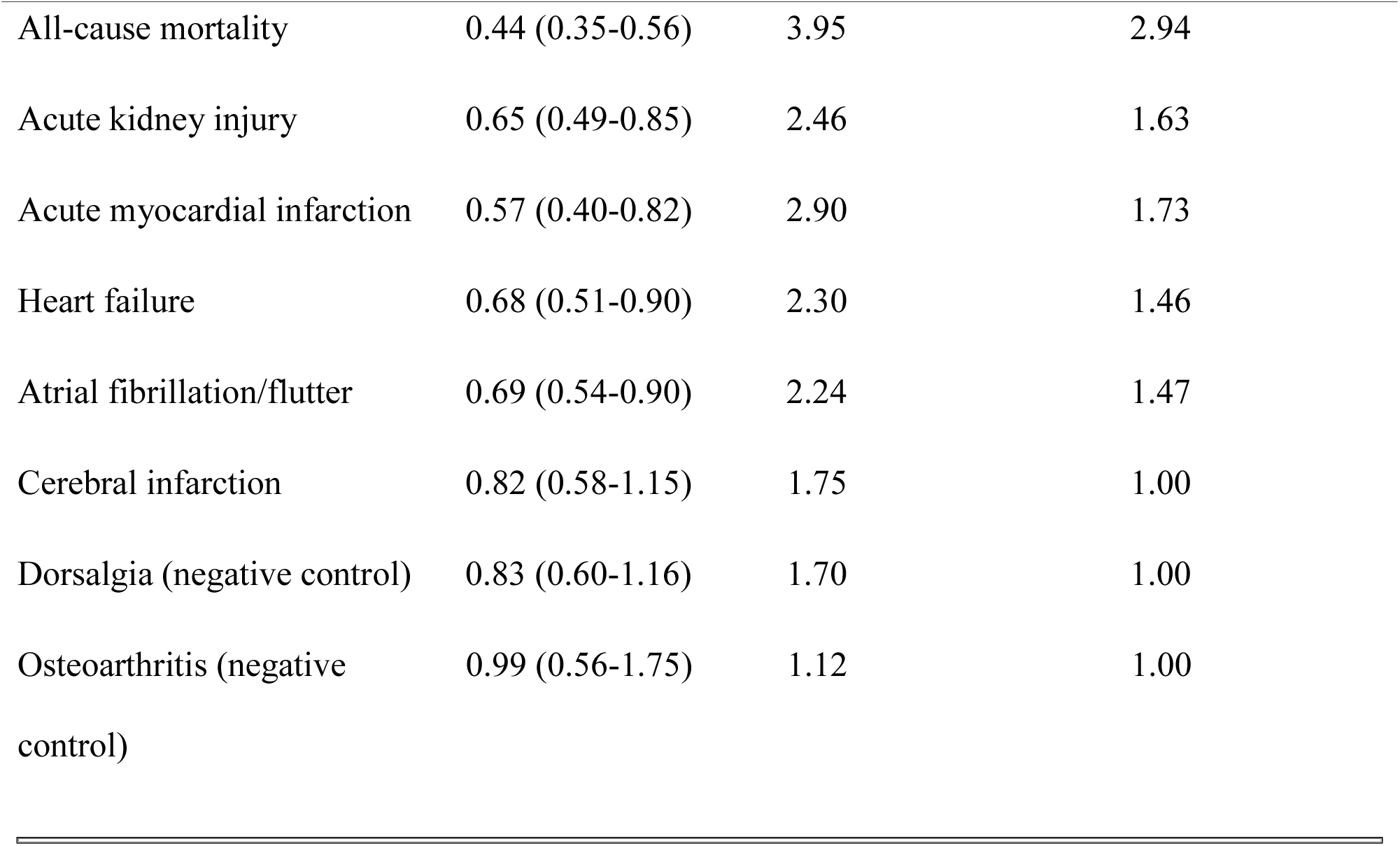
E-values for Primary 1-Year Outcomes.

| Outcome | Hazard Ratio (95%<br>CI) | E-value (Point<br>Estimate) | E-value (CI<br>Bound) |
| --- | --- | --- | --- |
| All-cause mortality | 0.44 (0.35-0.56) | 3.95 | 2.94 |
| Acute kidney injury | 0.65 (0.49-0.85) | 2.46 | 1.63 |
| Acute myocardial infarction | 0.57 (0.40-0.82) | 2.90 | 1.73 |
| Heart failure | 0.68 (0.51-0.90) | 2.30 | 1.46 |
| Atrial fibrillation/flutter | 0.69 (0.54-0.90) | 2.24 | 1.47 |
| Cerebral infarction | 0.82 (0.58-1.15) | 1.75 | 1.00 |
| Dorsalgia (negative control) | 0.83 (0.60-1.16) | 1.70 | 1.00 |
| Osteoarthritis (negative<br>control) | 0.99 (0.56-1.75) | 1.12 | 1.00 |

### Landmark Sensitivity Analysis

After exclusion of outcomes occurring within the first 30 days following SAVR, follow-up was assessed from day 31 through day 365 (Table 4). Associations with all-cause mortality (HR, 0.47; 95% CI, 0.34-0.66; P<.001), heart failure (HR, 0.65; 95% CI, 0.44-0.96; P=.030), and acute kidney injury (HR, 0.65; 95% CI, 0.44-0.95; P=.026) remained statistically significant, although heart failure and acute kidney injury associations were of borderline significance. By contrast, associations for acute myocardial infarction (HR, 1.05; 95% CI, 0.63-1.76; P=.85) and atrial fibrillation or flutter (HR, 0.84; 95% CI, 0.55-1.29; P=.43) lost statistical significance entirely, with the myocardial infarction point estimate moving slightly above 1.0. This pattern indicates that the myocardial infarction and atrial fibrillation signals observed in the primary analysis were predominantly driven by events occurring in the first 30 postoperative days. Cerebral infarction (HR, 0.97) and both negative-control outcomes (dorsalgia HR, 0.88; osteoarthritis HR, 0.85) remained null. The proportional hazards assumption was satisfied for heart failure in the landmark analysis (P=.004 result reflects rejection at landmark; visual inspection of curves did not suggest marked deviation).

**Table 4.**
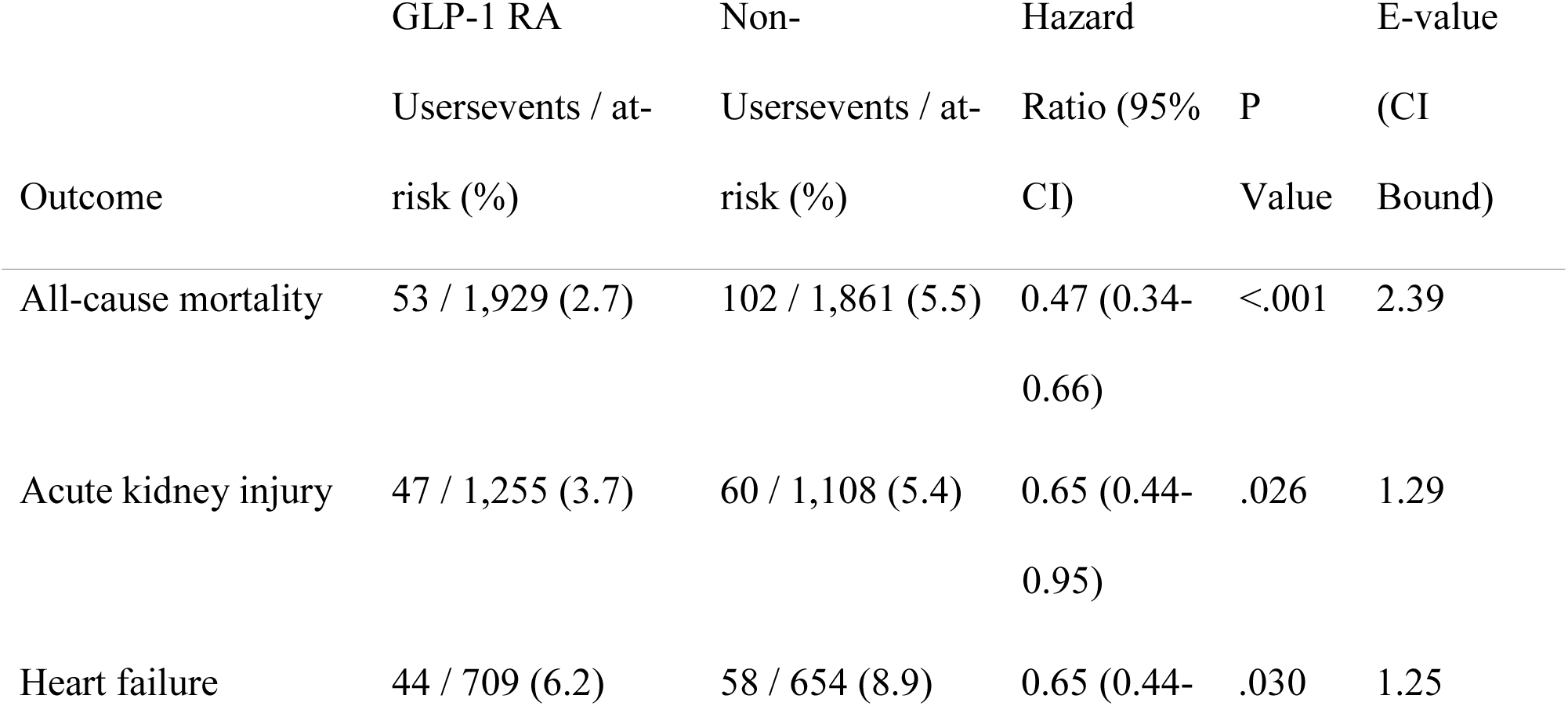
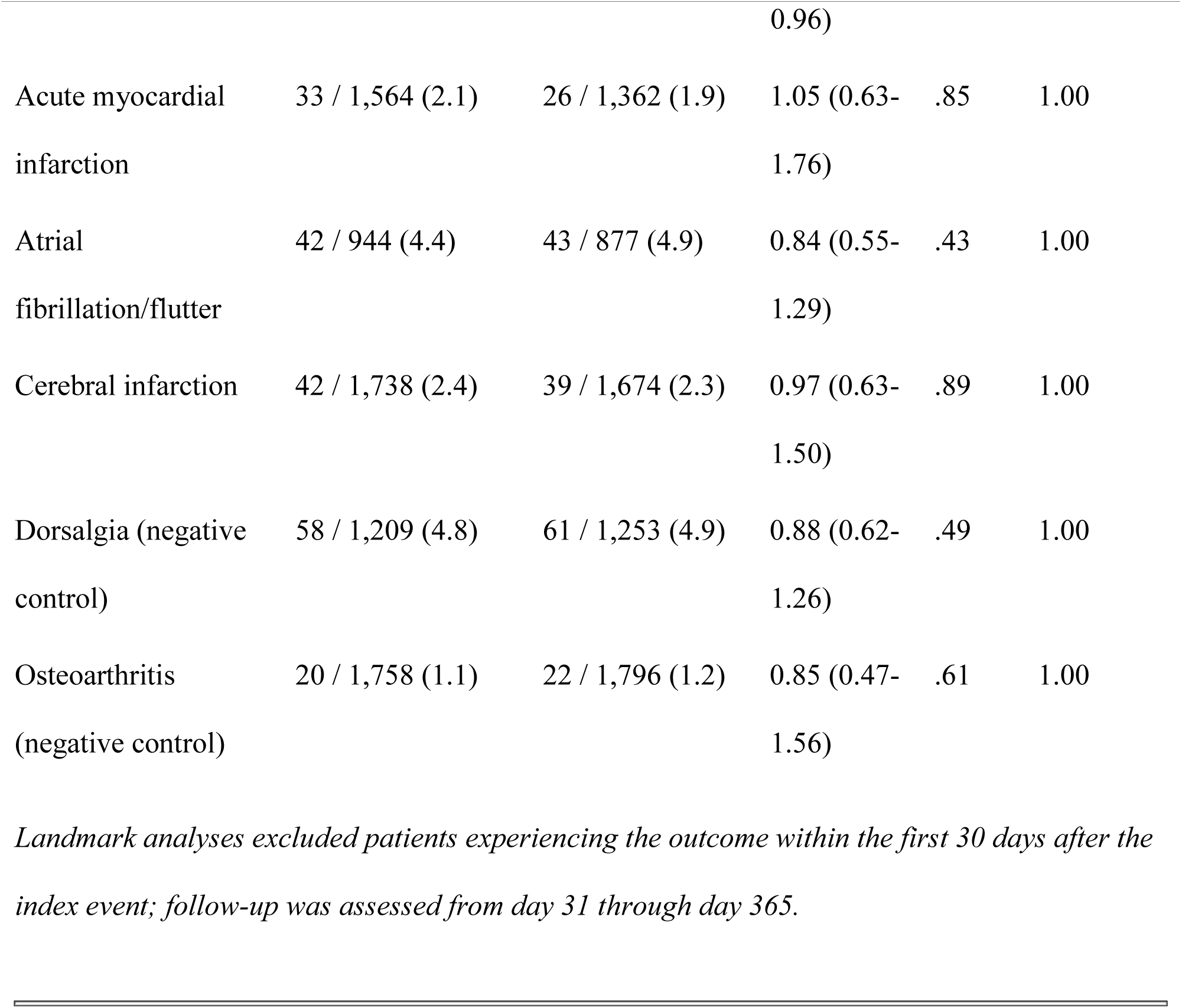
30-Day Landmark Sensitivity Analysis.

## Discussion

In this propensity-matched analysis of 3,968 adults with type 2 diabetes or obesity undergoing SAVR, perioperative GLP-1 RA use was associated with significantly lower 1-year risks of all-cause mortality, heart failure, acute kidney injury, acute myocardial infarction, and new-onset atrial fibrillation. Cerebral infarction did not differ between groups. Both prespecified negative-control analyses (dorsalgia and osteoarthritis) were null. In a 30-day landmark sensitivity analysis, associations for mortality, heart failure, and acute kidney injury persisted, while associations for myocardial infarction and atrial fibrillation became entirely null, indicating that these latter signals were predominantly driven by peri-procedural events.

### Context Within the Aortic Valve Intervention Literature

To our knowledge, this is the first analysis of GLP-1 RA use and cardiovascular outcomes in the SAVR population. Goyal et al^16^ reported a 1-year MACE hazard ratio of 0.63 (95% CI, 0.57-0.70) among 1,708 propensity-matched TAVR patients with diabetes or obesity, with directionally consistent associations across mortality, myocardial infarction, stroke, heart failure, and atrial fibrillation. Pham et al^17^ reported similar associations in a TriNetX-based analysis of TAVR patients with type 2 diabetes initiating GLP-1 RA therapy within 14 days of the procedure. The directional concordance between these TAVR analyses and the present SAVR findings suggest that favorable associations between GLP-1 RA use and post-procedural cardiovascular outcomes may extend across the spectrum of aortic valve intervention in cardiometabolic high-risk populations.

Two differences from the TAVR literature warrant comment. First, in the present SAVR cohort the myocardial infarction signal observed in the primary analysis (HR, 0.57) became entirely null after landmark restriction (HR, 1.05). This pattern was less pronounced in the Goyal et al TAVR analysis, in which myocardial infarction became non-significant after landmark restriction but remained directionally protective (HR, 0.74). The more complete attenuation in SAVR is likely to reflect the higher rate of peri-procedural myocardial injury associated with open cardiac surgery, cardiopulmonary bypass, and concomitant procedures compared with the transcatheter approach. Second, cerebral infarction was null in both our primary and landmark analyses, in contrast to the modest reduction observed by Goyal et al; this may reflect lower baseline stroke risk in the younger SAVR population or differences in stroke ascertainment between procedural contexts.

### Interpretation of Effect Magnitudes

The mortality association observed here (primary HR, 0.44; landmark HR, 0.47) is approximately double the effect observed in randomized cardiovascular outcome trials of GLP-1 RAs, in which hazard ratios for cardiovascular death have ranged from 0.78 to 0.91.^10–13^ This discrepancy is most plausibly driven, in substantial part, by residual confounding from frailty, functional status, and overall cardiometabolic care quality that propensity matching on EHR-extractable covariates cannot capture, rather than by GLP-1 RA pharmacology alone. The persistence of the mortality association after landmark restriction (and only modest attenuation from HR 0.44 to 0.47) is itself consistent with persistent selection effects: patients who survive 30 days after SAVR are those whose underlying care trajectory was already favorable, and GLP-1 RA exposure within this surviving subset may continue to mark this favorable trajectory in addition to any pharmacologic effect.

The SAVR population is a high-risk cardiometabolic cohort with elevated absolute event rates in which larger relative risk reductions are biologically plausible, and a true cardioprotective effect of GLP-1 RAs likely contributes to the observed associations. However, the observed magnitudes substantially overstate the causal effect attributable to drug pharmacology, and these estimates should be interpreted accordingly.

### Landmark Analysis and Biological Plausibility

The differential pattern in the landmark analysis has a coherent biological interpretation. The persistence of mortality, heart failure, and acute kidney injury associations after exclusion of the first 30 postoperative days is consistent with drug effects operating over a multi-month timescale through mechanisms such as weight loss, glycemic improvement, natriuresis, anti-inflammatory effects, and improved endothelial function,^14,15^ as well as with persistent confounding by care intensity and functional reserve. By contrast, the complete loss of the myocardial infarction signal (HR, 0.57 to 1.05) and the attenuation of the atrial fibrillation signal (HR, 0.69 to 0.84) are consistent with peri-procedural event capture. Post-cardiac-surgery atrial fibrillation peaks at 37% to 51% incidence within the first postoperative week after SAVR and is driven by surgical inflammation, atrial trauma, electrolyte shifts, and pericardial effusion,^8,9^ none of which would be expected to respond to GLP-1 RA therapy on a 30-day timescale. Similarly, peri-procedural myocardial injury after SAVR reflects ischemia-reperfusion physiology intrinsic to the procedure.

### Sensitivity to Unmeasured Confounding

E-values for the surviving landmark associations were 2.39 for mortality, 1.29 for acute kidney injury, and 1.25 for heart failure at the confidence interval bound. The mortality E-value indicates that an unmeasured confounder would need to be associated with both GLP-1 RA exposure and mortality by a risk ratio exceeding 2.39 to fully explain the association. The more modest E-values for acute kidney injury and heart failure indicate that confounders of relatively moderate strength could plausibly nullify these associations.

E-values quantify the minimum confounding strength required to nullify an association, not whether such confounding exists. Frailty is a dominant predictor of post-SAVR outcomes: a recent meta-analysis of cardiac surgical populations reported that frailty was associated with 3.6-fold increased odds of 30-day mortality (95% CI, 2.16-5.93) and 2.3-fold increased odds of 1-year mortality (95% CI, 1.56-3.25).^20^ Although frailty proxies were included among matching variables, the sensitivity of these administrative codes for true frailty is limited, and a frailty-related confounder of the magnitudes reported in cardiac surgical literature could substantially attenuate the observed associations.

### Competing Risk and Differential Mortality

A specific methodological concern relates to the differential mortality between cohorts and its impact on non-mortality outcome estimation. With a mortality HR of 0.44, GLP-1 RA users experienced approximately half the rate of death during follow-up compared with non-users. Because deaths were censored rather than treated as competing events for non-mortality outcomes, GLP-1 RA users had longer effective follow-up time during which non-fatal events could accrue, while non-users had more deaths censoring them out of the at-risk pool before non-fatal events could be observed. This censoring asymmetry systematically biases non-mortality hazard ratios in favor of GLP-1 RA users, meaning that the observed associations for heart failure, acute kidney injury, myocardial infarction, and atrial fibrillation are likely modestly stronger than the true subdistribution effects would be. The magnitude of this bias depends on the rate of death relative to the non-fatal outcome and on the correlation between death and the competing outcome; for outcomes strongly correlated with mortality such as heart failure, the bias is expected to be largest. Fine-Gray subdistribution hazard modeling, which would address this concern by treating death as a competing event, is not currently supported within the TriNetX platform; future analyses using patient-level extracts should incorporate competing-risk methods.

### Limitations

These findings should be interpreted as hypothesis-generating rather than evidence of causal benefit, and the limitations described below substantially constrain the interpretation of the observed associations.

Perhaps most importantly, patients receiving GLP-1 RAs likely represent a population with more intensive overall cardiometabolic management, including closer endocrinologic follow-up, more aggressive glycemic and weight optimization, and greater engagement with preventive care. The observed associations may therefore reflect the cumulative benefit of comprehensive metabolic management rather than an isolated GLP-1 RA drug effect, a distinction that propensity score matching on individual covariates cannot fully resolve. The magnitude of the mortality association (HR, 0.44, approximately double that observed in randomized trials) and the persistence of this association after landmark restriction together suggest that residual confounding from care quality and unmeasured cardiometabolic risk factors contributes substantially to the observed associations.

The exposure window (3 months before through 1 year after SAVR) is broad and encompasses heterogeneous clinical scenarios, including patients on established long-term GLP-1 RA therapy prior to surgery, patients initiated in the immediate postoperative period, and patients started months after discharge. These subgroups differ in biological plausibility of drug effect, duration of exposure, and susceptibility to survivorship bias, yet they are analyzed as a single exposed cohort. This temporal heterogeneity limits the ability to identify which phase of GLP-1 RA exposure, if any, drives the observed associations.

This observational analysis is also subject to confounding by indication; patients prescribed GLP-1 RAs were selected based on cardiometabolic indications, adequate functional status, and access to specialty pharmacy benefits, factors incompletely captured by matched covariates. Although frailty proxies (age-related debility, gait abnormalities, cognitive symptoms, cachexia) were included in matching, the sensitivity of these administrative codes for capturing true frailty is limited.

Residual baseline imbalances persisted after matching for body mass index (SMD, 0.30), hemoglobin A1c (SMD, 0.22), creatinine (SMD, 0.19), and estimated glomerular filtration rate (SMD, 0.11). The BMI and HbA1c imbalances are most plausibly interpreted as residual confounding by indication, as these characteristics directly informed the decision to prescribe GLP-1 RAs and likely capture broader differences in cardiometabolic care intensity not fully reflected in the matched covariates. The direction of bias introduced by each imbalance differs across outcomes and warrants explicit consideration. For all-cause mortality, the higher baseline BMI and HbA1c in the GLP-1 RA group would be expected to bias the mortality association toward the null, because these characteristics predict worse mortality independent of treatment; the observed protective association may therefore be a conservative estimate with respect to these specific confounders, although this attenuating effect is likely outweighed by the unmeasured residual confounding from frailty and care intensity discussed above. For heart failure, the higher BMI and HbA1c in the GLP-1 RA group should similarly bias toward the null given the established adverse prognostic effects of obesity and uncontrolled glycemia on incident heart failure. For acute kidney injury, the better kidney function in the GLP-1 RA group at baseline (lower creatinine, higher eGFR) likely contributes meaningfully to the observed lower risk because baseline kidney function is a strong predictor of postoperative AKI; the AKI association is therefore the outcome most vulnerable to this specific imbalance, consistent with its borderline statistical significance in the landmark analysis. For acute myocardial infarction and atrial fibrillation, both of which lost statistical significance in the landmark analysis, the residual imbalances are unlikely to substantially affect interpretation given the dominant role of peri-procedural mechanisms. For cerebral infarction, no significant association was observed; the residual imbalances do not change this null conclusion. A post-hoc multivariable-adjusted Cox model incorporating BMI, HbA1c, creatinine, and eGFR as additional covariates on top of the matched cohort would help quantify the residual confounding contribution and is identified as a future direction.

The exposure definition includes both prevalent users and postoperative initiators, conflating incident and prevalent drug effects. The prevalent-user design carries potential bias from depletion of susceptibles: patients who experienced adverse effects of GLP-1 RAs (intolerance, pancreatitis, gastrointestinal symptoms leading to discontinuation, or, hypothetically, early cardiovascular harm) are systematically removed from the exposed cohort before SAVR, leaving a population enriched for tolerance and benefit. This selection effect may contribute to the magnitude of the observed associations.

TriNetX does not reliably capture medication adherence, dosing, or duration of therapy. Outcomes were ascertained from EHR documentation within participating organizations, and events occurring outside the network, including out-of-hospital deaths, may be incompletely captured. The null negative-control outcomes (dorsalgia and osteoarthritis) argue against generalized healthy-user bias but cannot exclude outcome-specific confounding. The osteoarthritis association was statistically null (HR, 0.99) but its wide confidence interval (0.56-1.75) does not exclude clinically meaningful effects in either direction.

Formal competing-risk analysis was not performed; deaths were censored rather than treated as competing events, biasing non-mortality hazard ratios in favor of GLP-1 RA users as discussed above. The SAVR population in TriNetX is heterogeneous, encompassing isolated SAVR and SAVR with concomitant procedures; subgroup analyses by surgical complexity were not performed in this initial analysis.

### Future Directions

Future analyses should stratify by timing of GLP-1 RA initiation relative to surgery, comparing patients with established presurgical use, those initiated in the immediate postoperative period and those started in the late postoperative phase. This stratification would clarify whether the observed associations are driven by chronic preoperative metabolic optimization, acute perioperative drug effects, or post-discharge cardiometabolic management. Additional prespecified subgroup analyses stratified by diabetes status, obesity status, and isolated versus concomitant SAVR would clarify whether the observed associations vary by metabolic phenotype and surgical complexity. An active comparator analysis comparing GLP-1 RA initiators to new users of DPP-4 inhibitors or SGLT2 inhibitors would further address confounding by indication and survivorship bias. A post-hoc multivariable-adjusted Cox model including the residually imbalanced covariates (BMI, HbA1c, creatinine, eGFR) as additional adjustments on top of the matched cohort would help quantify the contribution of these imbalances to the observed associations. Fine-Gray subdistribution hazard modeling using patient-level data extracts would address the competing risk of death for non-mortality outcomes. External validation in an independent data source would strengthen generalizability. Ultimately, prospective randomized evaluation is required to determine whether GLP-1 RA therapy confers genuine cardiovascular benefit in patients undergoing SAVR.

### Conclusions

In this propensity-matched analysis of adults with type 2 diabetes or obesity undergoing SAVR, perioperative GLP-1 RA use was associated with significantly lower 1-year risks of all-cause mortality, heart failure, acute kidney injury, acute myocardial infarction, and new-onset atrial fibrillation. Associations with mortality, heart failure, and acute kidney injury persisted in a 30-day landmark sensitivity analysis. By contrast, associations for myocardial infarction and atrial fibrillation became entirely null after landmark restriction, indicating these signals predominantly reflected peri-procedural events. Negative-control analyses for both dorsalgia and osteoarthritis were null. The magnitude of observed associations exceeds randomized-trial estimates and likely reflects substantial residual confounding from frailty, functional status, and overall cardiometabolic care intensity rather than GLP-1 receptor agonist pharmacology alone.

These hypothesis-generating findings extend prior observations from the TAVR literature to the SAVR population and support prospective randomized investigation of perioperative GLP-1 RA therapy in patients undergoing surgical aortic valve replacement.

## Acknowledgments

Open Evidence was used to assist with formatting and editing portions of this manuscript. It was not used for study design, data acquisition, data analysis, or interpretation of results. The authors reviewed all content and take full responsibility for the accuracy, validity, and originality of the manuscript.

## Sources of Funding

None.

## Disclosures

None.

## Data Availability

The data underlying this study were obtained from the TriNetX U.S. Collaborative Network under a license agreement and are not publicly available. TriNetX data consist of de-identified electronic health record information aggregated from participating healthcare organizations; restrictions imposed by the data use agreement prohibit the authors from redistributing patient-level data. Researchers may request access to the TriNetX platform directly from TriNetX (https://trinetx.com). The cohort definitions, diagnostic and procedure codes, exposure and outcome specifications, covariates, and analytic methods used in this study are described in detail in the Methods to permit reproduction by investigators with TriNetX access. Additional information regarding the analytic approach is available from the corresponding author upon reasonable request.

---

**Figure 2.**
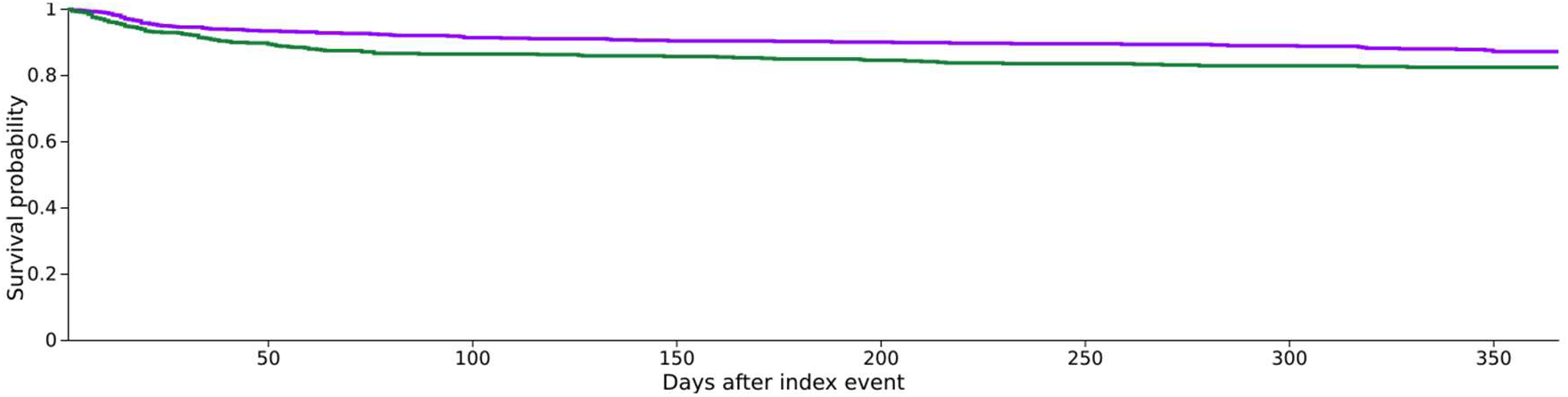
Kaplan-Meier curve for heart failure over 1-year follow-up after propensity score matching. Purple curve represents GLP-1 RA users; green curve represents non-users.

**Figure 3.**
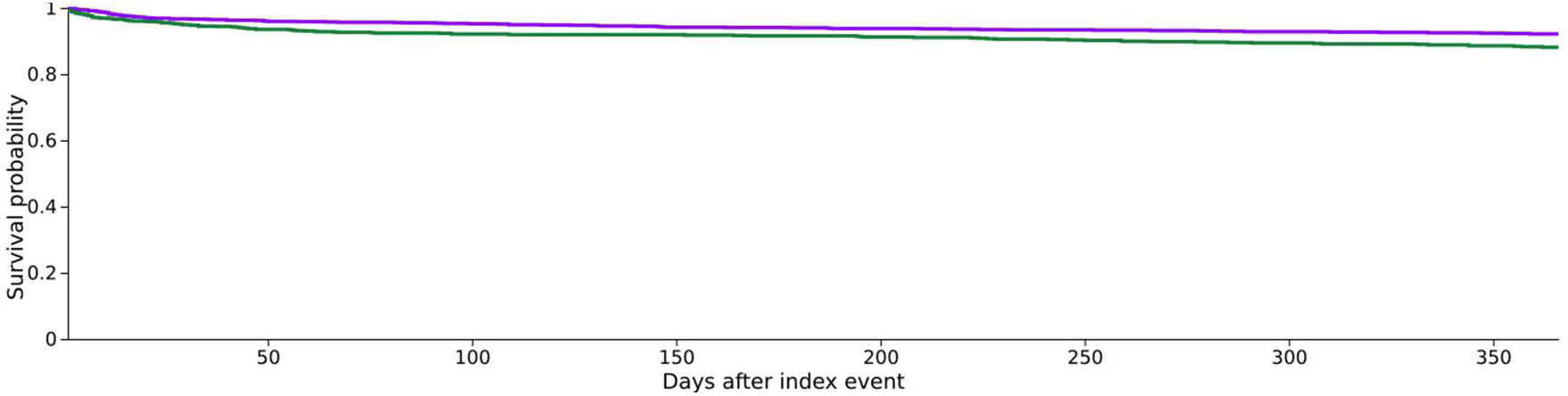
Kaplan-Meier curve for acute kidney injury over 1-year follow-up after propensity score matching. Purple curve represents GLP-1 RA users; green curve represents non-users.

**Figure 4.**
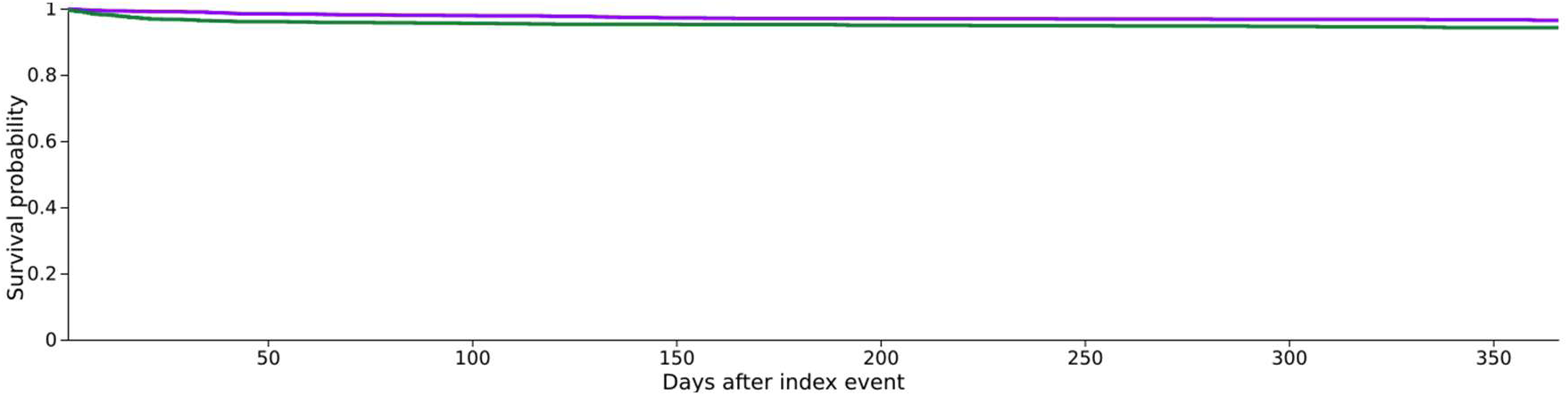
Kaplan-Meier curve for acute myocardial infarction over 1-year follow-up after propensity score matching. Purple curve represents GLP-1 RA users; green curve represents non-users.

**Figure 5.**
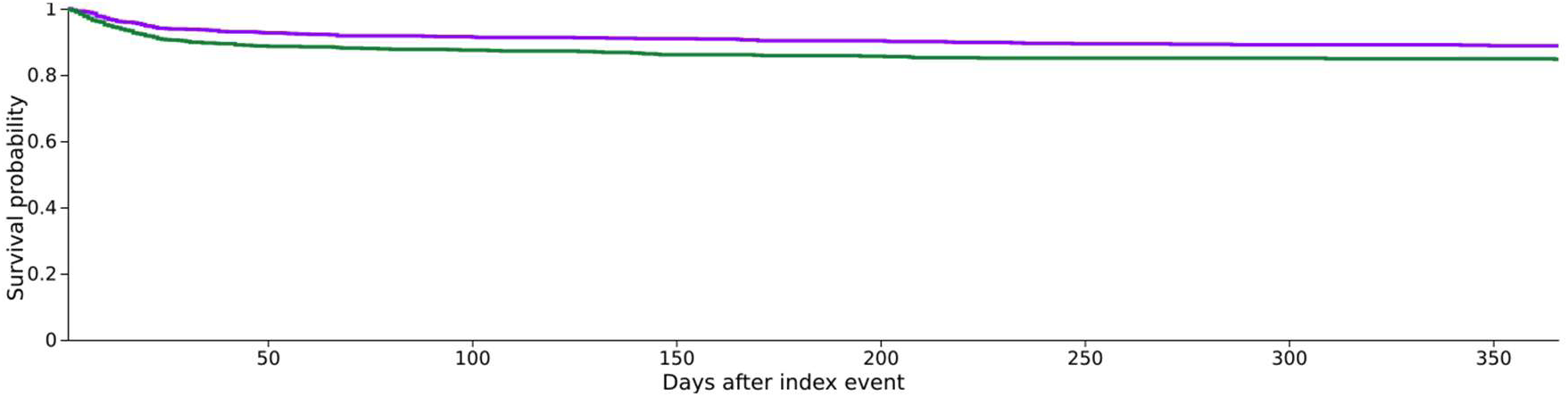
Kaplan-Meier curve for atrial fibrillation/flutter over 1-year follow-up after propensity score matching. Purple curve represents GLP-1 RA users; green curve represents non-users.

**Figure 6.**
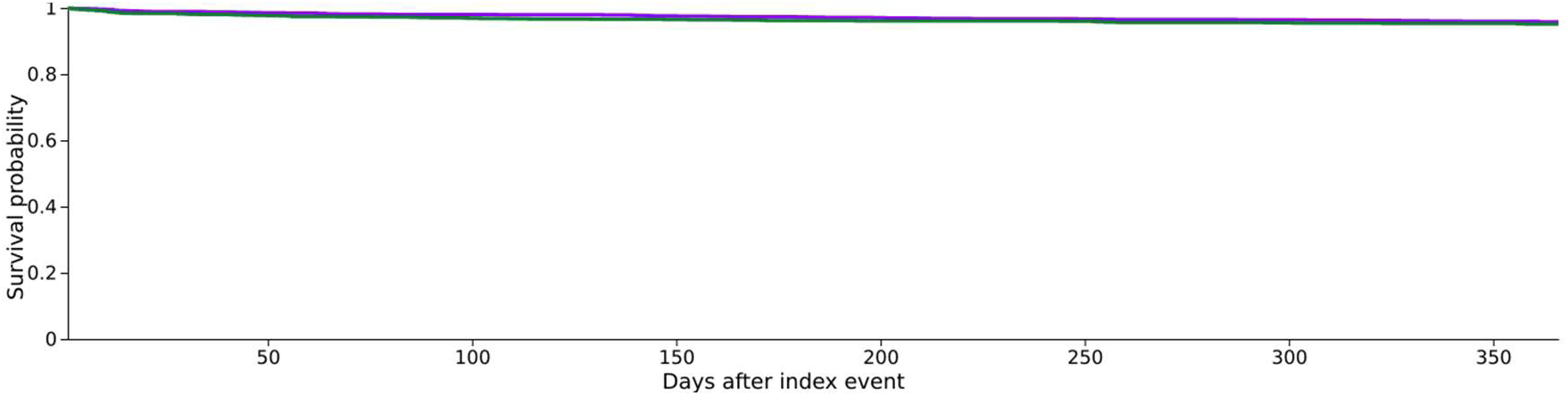
Kaplan-Meier curve for cerebral infarction over 1-year follow-up after propensity score matching. Purple curve represents GLP-1 RA users; green curve represents non-users.

**Figure 7.**
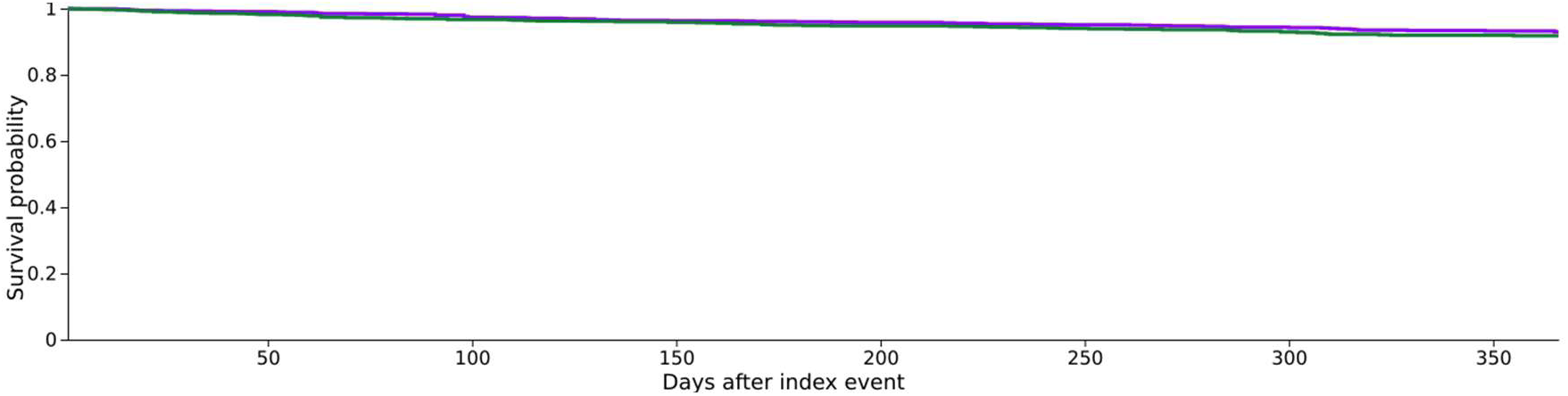
Kaplan-Meier curve for dorsalgia (negative-control outcome) over 1-year follow-up after propensity score matching. Purple curve represents GLP-1 RA users; green curve represents non-users.

**Figure 8.**
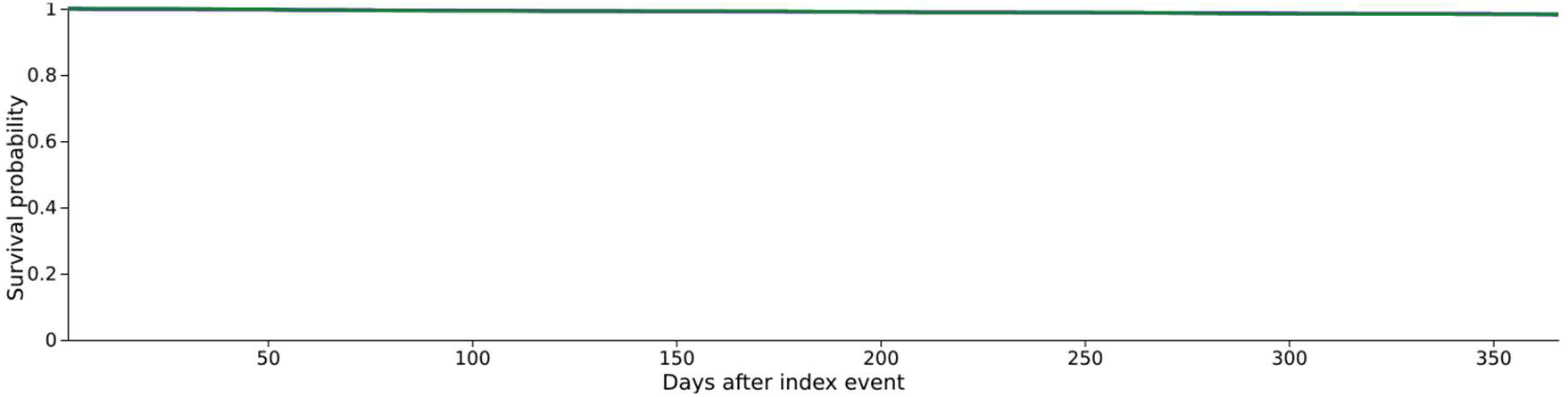
Kaplan-Meier curve for osteoarthritis (negative-control outcome) over 1-year follow-up after propensity score matching. Purple curve represents GLP-1 RA users; green curve represents non-users.

## Nonstandard Abbreviations and Acronyms

GLP-1 RA: glucagon-like peptide-1 receptor agonist
SAVR: surgical aortic valve replacement
TAVR: transcatheter aortic valve replacement
EHR: electronic health record
SMD: standardized mean difference
HR: hazard ratio

## Notes

### Competing Interest Statement

The authors have declared no competing interest.

### Funding Statement

No external funding was received.

### Author Declarations

This study used aggregated, de-identified data from the TriNetX federated research network and did not constitute human subjects research as defined under 45 CFR section 46.102. Because the analysis involved only de-identified data and no individually identifiable private information, it was exempt from institutional review board review under 45 CFR section 46.104(d)(4). Informed consent was not required.

